# Development and external validation of prediction models for major osteoporotic fracture and hip fracture in people with intellectual disability

**DOI:** 10.1101/2025.02.19.25322527

**Authors:** Margaret Smith, Jan Roast, Gary S. Collins, Tim A. Holt, Valeria Frighi

**Author notes:** Corresponding author Dr Tim A Holt.

## Abstract

**Purpose:** Compared with the general population, people with intellectual disabilities (ID) have higher incidences of major osteoporotic fracture (MOF) and hip fracture (HF), and osteoporosis develops at a younger age. The rate of HF in those aged 50 years and over is two and four times higher than that in women and men, respectively, without ID. It is essential to identify people with ID at risk of such fractures so that a targeted fracture prevention strategy can be designed. However, current fracture prediction models are derived from the general population and may underestimate risk in the ID population.

**Methods:** Prediction models (IDFracture) for the 10-year risk of HF and MOF were developed and validated in populations of people with ID aged 30-79 years. Models were developed in the CPRD GOLD database and temporally validated in the Aurum database. The predictors included those in current fracture prediction models and ID-specific predictors such as Down syndrome. All the predictors were included in the Cox regression models. Bootstrapping was used to adjust for overfitting.

**Results:** The development cohort included 38,665 people with IDs, 1045 with MOFs and 360 with HFs within 10 years. The external validation cohort included 76,385 people, 2420 MOFs and 1001 HFs. Discrimination, as judged by the C statistic, was good: MOF 0.775, HF 0.839. The calibration was also good but tended to overpredict at the highest predicted risks.

**Conclusion:** IDFracture has potential as a screening tool in clinical practice to identify people with ID who are at increased risk of MOF and HF.

**Mini Abstract:** *Brief rationale:* People with intellectual disability (ID) have a relatively high incidence of fracture, so current risk prediction models are not appropriate.

*Main result:* A new prediction model for people with ID showed good calibration and discrimination in external validation.

*Significance of paper:* This is the first such prediction model developed for people with ID.

## Introduction

Major epidemiological studies have shown that people with intellectual disabilities (ID) have a higher incidence of fracture than their general population counterparts [1-4]. In the most recent and comprehensive of these studies, the difference was particularly evident for major osteoporotic fractures (MOFs), especially for hip fracture (HF) [3]. Among those aged 50 years and over, women with ID had a 2·3 times higher rate and men had a 3·8 times higher rate of HF than women and men without ID. Moreover, people with ID start to develop osteoporotic fractures at a younger age than in the general population. These findings clearly highlight the need for fracture prevention strategies in individuals with ID. However, for any such strategies to be effective, it is necessary to identify those at increased risk. In the general population, the fracture prediction models FRAX and QFracture are widely used to identify individuals at increased risk of MOF and HF [5, 6]. Neither of these models include a diagnosis of ID as a predictor, and given the differences in fracture incidence, the available fracture prediction models may not provide accurate estimates in individuals with ID. Individuals with ID have accompanying clinical characteristics, e.g., syndromal disorders associated with medical complications, which are different or more frequent than in the rest of the population. These individuals may have additional predictors of fracture, which could be incorporated into a fracture prediction model developed for this population.

### Purpose

In this paper, we describe the derivation and external validation of two prediction models (IDFracture) to estimate the 10-year risk of MOF and HF in individuals with ID.

## Methods

The prediction models were derived and internally validated in a cohort study using data extracted from the Clinical Practice Research Datalink (CPRD) GOLD database along with linked Hospital Episode Statistics (HES) inpatient data (NHS digital) for England. CPRD GOLD is a database of anonymised primary care records derived from practices in the United Kingdom that use the Vision software system [7]. External validation was performed using a cohort extracted from the CPRD Aurum database along with linked HES data. The Aurum database became available after the project started and contains anonymised records from practices that use the EMIS software system [8].

The code lists that we used for this study are published on our GitHub site (*link to be added*). The reporting of this study followed the Transparent Reporting of a prediction model for Individual Prognosis Or Diagnosis (TRIPOD+AI) statement [9]. Data management and analyses were performed in Stata 16 [10].

### Study populations

The derivation cohort was a subset of the cohort used for a study of fracture incidence in individuals with ID in CPRD GOLD [3]. The records of all individuals with a code indicating an ID who were registered at practices in England during the study period (1/1/1998 to 31/12/2017) and linked to the HES Admitted Patient Care database were extracted. The data for each included individual also had to be “acceptable”, meaning that the patient was permanently registered and that data on key variables had passed basic quality checks. We used the code list of the Quality and Outcomes Framework Learning Disability Register as defined in their business rules. We included diagnostic and service user codes [3]. General learning difficulty codes were excluded if accompanied by one or more codes for specific learning difficulties (e.g. dyslexia). We also excluded codes for ID if they were observed during pregnancy (i.e. may have resulted from genetic screening of a foetus), and we excluded codes for some sex-linked syndromes in females. For each patient, the index date was defined as the latest date of 1/1/1998, contributing 365 days of “up to standard” (UTS) data in their current practice and reaching the age of 30 years. The UTS date is the date at which the practice is considered to have continuous high-quality data suitable for use in research, on the basis of an algorithm that looks at gaps in the data and practices death recording. Individuals older than 79 years of age at index were excluded because of the small number of individuals in this age group.

The cohort for external validation consisted of people with ID in the Aurum database. To evaluate the performance of the prediction models during a more recent study period, records of individuals with codes indicating ID, “acceptable” data available in CPRD Aurum between 1/1/2008 and 31/10/2020, and linked to HES inpatient data were extracted. Aurum does not contain an “up to standard” date, so the index date was the latest of 1/1/2008, contributing 365 days of data in their current practice and reaching the age of 30 years. Practices that had previously contributed to the GOLD database were excluded from the validation cohort. We used the same definition of ID as for the GOLD cohort except that we omitted service user codes from the Aurum population. We did this because the number of people with only these codes was much greater than in the GOLD derivation cohort.

### Outcomes

In both the derivation cohort and the external validation cohort, the follow-up period was a maximum of 10 years from the index date or until the earliest of the outcome date, the study end date, the date of leaving the current practice due to death or other reasons, and the last date of download of data from their practice. The outcomes to be predicted were either any MOF or a HF within 10 years of the index date. MOFs and HFs were identified from primary care and from linked HES data [3]. Owing to the difficulty in determining the mechanism of fracture from patients’ clinical records, the MOF was defined by the anatomical sites that are generally affected. Consistent with current definitions and previous studies, MOF sites include the hip, wrist, vertebra and shoulder and unspecified sites [11]. The outcome date was the date of the first code for HF or MOF after the index date.

### Predictors

The variables included in the models were predictors included in QFracture or FRAX and other potential predictors of fracture in individuals with ID. The latter group of predictors was chosen on the basis of the literature, our own clinical experience, an association with low bone mineral density, or the high prevalence in the ID population combined with their potentially relevant clinical significance. Predictor variables were extracted from records prior to the index date. The exception was the previous fracture, where linked HES data were also used. To prevent double-counting of fractures as both predictors and outcomes, fractures occurring within 6 months after the index date were not counted as outcomes if another fracture at the same site was recorded within the previous 6 months and prior to the index date.

We included all variables from the general population fracture prediction models except for family history of osteoporosis or hip fracture and personal history of living in a nursing or care home. These variables were dropped because information on family history would have been missing for a large proportion of patients and because the living arrangements for individuals with ID differ from those without ID and relate more to social than to health factors. We did not distinguish between type 1 diabetes and type 2 diabetes, as the coding is not always clear. As our study population was much smaller than that of the QFracture study, we combined the levels of some categorical variables, e.g., smoking was included as a binary variable (current smoker or nonsmoker) [6].

We included additional variables that might be predictors of fracture in the ID population on the basis of clinician experience and a literature review. These included height (cm), impaired mobility (yes/no), hearing impairment (yes/no), self-injurious behaviour (yes/no), visual impairment (yes/no), a prescription for sedatives or hypnotics (yes/no), a prescription for proton pump inhibitors (yes/no), a prescription for antipsychotics (yes/no), a prescription for risperidone and/or hyperprolactinaemia (yes/no), type of ID, severity of ID, and hypogonadism (yes/no). The type of ID was coded as Down syndrome, other specified genetic or congenital syndromes, or other/unknown. The severity of the ID was coded as mild if there was a code specifying this; otherwise, it was classified as not mild. Hypogonadism (which also includes long-term progesterone treatment in women) was included because of its profound effect on bone and its association with ID as a potential comorbid condition and as an iatrogenic complication [12-15]. Vitamin D deficiency could not be included as a potential predictor because of poor and inconsistent recording of vitamin D levels in the database [16, 17].

### Missing data

Individuals with missing smoking data were assumed to be nonsmokers; those with missing alcohol data were assumed not to be moderate/heavy drinkers; and those with missing ethnicity data were coded as white. Missing values of body mass index (BMI) and height were imputed with multiple imputation by chained equations, predictive mean matching was used, and all variables to be included in the planned analyses (including the outcome) were included. Thirty imputed datasets were obtained for each outcome. The same sets of imputations were used for all analyses in each cohort.

### Model derivation

The models were developed and internally validated in the cohort extracted from the GOLD database. We calculated descriptive statistics for baseline characteristics. We also plotted Kaplan–Meier curves for hip or osteoporotic fractures. We used Cox proportional hazard models to estimate hazard ratios (HRs). The proportional hazards assumption was checked graphically. The crude HR and HR adjusted for sex and age (as a continuous covariate) were calculated for HF or MOF.

There were no or very few HF outcomes in the presence of some binary predictors (e.g., alcohol consumption, malabsorption). These predictors were excluded from the HF models but included in the MOF models. Liver disease and chronic kidney disease were also combined into a single variable (liver or kidney disease) in the HF model. The multivariable models included all remaining predictors. Fractional polynomials were used to explore any nonlinearity for the three continuous variables (age, BMI and height) via the Stata mfpmi module [18]. Untransformed age, BMI and height were selected for both the HF and MOF models. Interaction terms for age and sex were included in the final prediction models. Adjusted HRs were then calculated in models containing all the predictors and interaction terms. The predicted risk for each individual was then calculated from the log hazard ratios and the 10-year baseline survival estimate from the multivariable model [19].

### Internal validation

Calibration plots were plotted via the pmcalplot after combining predicted risks over imputations [20]. We also assessed the apparent model performance with Harrell’s C and Royston and Sauerbrei’s R^2^_D_ and D statistics [21, 22]. Harrell’s C is the probability that within a randomly selected pair of patients, a patient with a shorter survival time has a higher predicted risk. The D statistic is an estimate of the log HR of the outcome comparing two equal-sized prognostic groups. R^2^_D_ is a transformation of the D statistic that can be interpreted as the proportion of variation in the outcome that is explained by the model. The performance statistics were calculated separately for each imputed dataset and then combined via Rubin’s rules.

We used bootstrapping to correct the validation statistics for optimism as a result of overfitting. One thousand bootstrap samples were taken from the imputed data. The model including all covariates was fitted in each bootstrap sample, replicating the model-building strategy in the development cohort [19]. Approximate confidence intervals were estimated by shifting the location of the 2·5th and 97·5th percentiles of the distribution of validation statistics on the bootstrap samples [23]. For each outcome, a uniform shrinkage coefficient for the calibration slope was estimated via the same bootstrapping method used for the calibration slope. The shrinkage coefficient was applied to the linear predictor, and the 10-year baseline survival was re-estimated to produce the final prediction models.

### External validation

We calculated descriptive statistics and univariate, age- and sex-adjusted HRs for the Aurum validation cohort as described above. The ten-year predicted risk of fracture was estimated by applying the final prediction models to this cohort. Calibration plots and validation statistics were then estimated for the entire cohort. Approximate confidence intervals were estimated from the distribution of these statistics in 600 bootstrapped samples. Further calibration plots and validation statistics were calculated on subgroups of age, sex, Index of Multiple Deprivation and index year.

## Results

The GOLD derivation cohort included 38,665 people, 43.1% of whom were female (Table 1). The median age at the index date was 38.9 years. The percentage of patients with Down syndrome was 10.1%, and an additional 5.1% had another specified type of ID. Only 3.3% had a record specifically stating mild ID. The prevalence of MOF prior to the index was 6.2%, but the prevalence of fracture at other sites was 15.5%. The prevalence of epilepsy (diagnosis or prescription of anticonvulsants) was 20.6%. There were 360 HF and 1045 MOF fractures in this population over a median of 6.3 years of follow-up. The ten-year cumulative incidences of HF and MOF were 1.6% and 4.6%, respectively (Figure S1 and S2). The Aurum validation population included 76,385 people with IDs, 40.9% of whom were female. The median age was 41.5 years (Table 1). There were slightly fewer people with previous MOF fractures (5.5%), but the proportions diagnosed with comorbidities or prescribed medications were generally higher than in the GOLD cohort. The percentage of patients with Down syndrome was 7.9%, and an additional 6.3% had another specified type of ID. A total of 11.2% had a record specifically stating mild ID. There were 1001 HF and 2420 MOF fractures with a median follow-up of 4.8 years. The ten-year cumulative incidences of HF and MOF were higher than those in the derivation cohort (2.5% and 6.1%, respectively).

**Table 1.** Descriptive statistics for the derivation and validation cohorts of people with intellectual disability.

|  | Derivation cohort |  | Validation cohort |  |
| --- | --- | --- | --- | --- |
| N | 38,665 | (100%) | 76,385 | (100%) |
| Index year (median, 10-90 percentiles) | 2004 | (1998-2014) | 2011 | (2008-2019) |
| Age at index (years) (median, 10-90 percentiles) | 38.9 | (30.0-61.0) | 41.5 | (30.0-63.5) |
| Female | 16,663 | (43.1%) | 31,206 | (40.9%) |
| Ethnicity |  |  |  |  |
| Non-white | 1,061 | (2.7%) | 7,772 | (10.2%) |
| White | 21,415 | (55.4%) | 63,103 | (82.6%) |
| Missing | 16,189 | (41.9%) | 5,510 | (7.2%) |
| Body mass index (kg/m<sup>2</sup>) (median, 10-90 percentiles) | 26.7 | (20.1-36.3) | 27.7 | (20.4-38.5) |
| Missing body mass index | 13,501 | (34.9%) | 21,597 | (28.3%) |
| Height (cm) (median, 10-90 percentiles) | 165 | (152-181) | 167 | (151-182) |
| Missing height | 14,019 | (36.3%) | 18,992 | (24.9%) |
| Current smoker | 7,430 | (19.2%) | 16,164 | (21.2%) |
| Non smoker /ex smoker | 23,061 | (59.6%) | 57,565 | (75.4%) |
| Missing smoking status | 8,174 | (21.1%) | 2,656 | (3.5%) |
| Current moderate/ heavy drinker<sup>a</sup> | 4,511 | (11.7%) | 2,807 | (3.7%) |
| Non drinker/ light/ drinker/ ex drinker | 21,962 | (56.8%) | 24,852 | (32.5%) |
| Missing alcohol status | 12,192 | (31.5%) | 48,726 | (63.8%) |
| Previous major osteoporotic fracture (MOF) | 2,404 | (6.2%) | 4,229 | (5.5%) |
| Previous fracture at non-MOF site | 5,982 | (15.5%) | 17,162 | (22.5%) |
| Dementia | 326 | (0.8%) | 1,384 | (1.8%) |
| History of falls | 2,073 | (5.4%) | 6,148 | (8.0%) |
| Malabsorption | 336 | (0.9%) | 1,386 | (1.8%) |
| Endocrine problems | 326 | (0.8%) | 874 | (1.1%) |
| Asthma or chronic obstructive pulmonary disease | 4,692 | (12.1%) | 12,217 | (16.0%) |
| Cancer | 432 | (1.1%) | 1,696 | (2.2%) |
| Heart disease | 832 | (2.2%) | 2,540 | (3.3%) |
| Epilepsy or taking anticonvulsants | 7,982 | (20.6%) | 20,940 | (27.4%) |
| Rheumatoid arthritis or systemic lupus erythematosus | 154 | (0.4%) | 361 | (0.5%) |
| Liver disease | 332 | (0.9%) | 178 | (0.2%) |
| Chronic kidney disease | 314 | (0.8%) | 1,971 | (2.6%) |
| Diabetes | 1,988 | (5.1%) | 5,982 | (7.8%) |
| Steroid tablets | 276 | (0.7%) | 623 | (0.8%) |
| Oestrogen only hormone replacement therapy | 129 | (0.3%) | 205 | (0.3%) |
| Antidepressants | 5,640 | (14.6%) | 15,021 | (19.7%) |
| Antipsychotics | 5,387 | (13.9%) | 13,234 | (17.3%) |
| Poor mobility/Parkinsons | 3,875 | (10.0%) | 10,440 | (13.7%) |
| Hearing impairment | 4,036 | (10.4%) | 10,858 | (14.2%) |
| Self-injurious behaviour | 1,069 | (2.8%) | 1,926 | (2.5%) |
| Visual impairment | 1,881 | (4.9%) | 7,134 | (9.3%) |
| Hypogonadism (including taking progesterone) | 1,791 | (4.6%) | 4,080 | (5.3%) |
| Sedatives and hypnotics | 2,989 | (7.7%) | 6,078 | (8.0%) |
| Proton pump inhibitors | 2,737 | (7.1%) | 9,403 | (12.3%) |
| Risperidone and/or hyperprolactinaemia | 1,786 | (4.6%) | 4,948 | (6.5%) |
| Specified type of intellectual disability<sup>b</sup> | .. | .. | .. | .. |
| Down syndrome | 3,907 | (10.1%) | 6,069 | (7.9%) |
| Other specified intellectual disability | 1,977 | (5.1%) | 4,796 | (6.3%) |
| Mild severity of intellectual disability<sup>c</sup> | 1,286 | (3.3%) | 8,578 | (11.2%) |
| Follow-up (years) (median, 10-90 percentiles) | 6.3 | (0.8-10.0) | 4.8 | (0.6-10.0) |

Statistics are n (%) unless otherwise stated.
a Moderate/heavy drinking is defined as 3+ alcohol units daily
b The remainder of the cohort had an unspecified intellectual disability.
c The remainder of the cohort had non-mild or unspecified severity of intellectual disability.

The univariate and age- and sex-adjusted HRs for each of the predictors were generally similar for the derivation and validation cohorts, although confidence intervals for HRs from the derivation cohort were wide in place (Tables S1 and S2). The HRs for the derivation cohort, for the multivariate model containing all of the predictors, are given in Table 2. The HR for MOF was greater in women than in men at age 40, and the risk also increased more rapidly with age in women than in men. The HR for HF was similar in women and men at age 40, but again, it increased more with age in women. However, it more than doubled per ten years of age in both sexes. Other statistically significant predictors for MOF were moderate/heavy alcohol consumption, previous MOF, previous other fracture, history of falls, cancer, epilepsy, diabetes, treatment with antidepressants and hypogonadism, which were all associated with increased risk. A higher BMI predicted a lower risk of developing an MOF. Heart disease and poor mobility/Parkinson’s disease also appeared to be associated with a lower risk. Other predictors associated with a higher risk of HF were previous MOF, previous other fractures, history of falls, cancer, epilepsy, diabetes and Down syndrome. A higher BMI predicted a lower risk of HF.

**Table 2.** Hazard ratios (HR) (95% confidence intervals (CI)) for predictors of major osteoporotic fracture (MOF) and hip fracture (HF) in the derivation cohort. Hazard ratios were estimated via multivariate Cox regression models containing all predictor variables for MOF or HF.

| Predictor | MOF (n=1045) |  | HF (n=360) |  |
| --- | --- | --- | --- | --- |
|  | HR | (95% CI) | HR | (95% CI) |
| Ten years older age in females | 1.75 | (1.64-1.87) | 2.30 | (2.05-2.58) |
| Ten years older age in males | 1.56 | (1.45-1.67) | 2.08 | (1.86-2.33) |
| Female at age 40 years | 1.25 | (1.01-1.54) | 0.95 | (0.64-1.39) |
| Five units higher body mass index (kg/m<sup>2</sup>) | 0.86 | (0.81-0.92) | 0.71 | (0.62-0.80) |
| 10cm taller | 0.97 | (0.87-1.09) | 0.96 | (0.81-1.14) |
| Current smoker | 0.92 | (0.77-1.10) | 0.84 | (0.61-1.15) |
| Current moderate/ heavy drinker<sup>a</sup> | 1.28 | (1.05-1.56) | 1.09 | (0.75-1.60) |
| Non-white ethnicity | 0.65 | (0.36-1.19) | .. | .. |
| Previous major osteoporotic fracture (MOF) | 2.70 | (2.26-3.23) | 1.89 | (1.37-2.61) |
| Previous fracture at non-MOF site | 1.60 | (1.36-1.89) | 1.45 | (1.09-1.93) |
| Dementia | 1.31 | (0.83-2.07) | 1.30 | (0.69-2.45) |
| History of falls | 1.65 | (1.35-2.01) | 1.74 | (1.25-2.41) |
| Malabsorption | 1.40 | (0.77-2.54) | .. | .. |
| Endocrine problems | 1.41 | (0.87-2.29) | .. | .. |
| Asthma or chronic obstructive pulmonary disease | 1.17 | (0.96-1.43) | 1.06 | (0.73-1.54) |
| Cancer | 1.49 | (1.00-2.21) | 1.77 | (0.99-3.18) |
| Heart disease | 0.69 | (0.48-0.99) | 0.66 | (0.37-1.16) |
| Epilepsy or taking anticonvulsants | 1.73 | (1.51-1.99) | 1.89 | (1.49-2.40) |
| Rheumatoid arthritis or systemic lupus erythematosus | 1.51 | (0.63-2.09) | .. | .. |
| Liver disease | 1.15 | (0.80-2.84) | .. | .. |
| Chronic kidney disease | 1.23 | (0.69-2.20) | .. | .. |
| Liver or kidney disease | .. | .. | 1.02 | (0.48-2.18) |
| Diabetes | 1.27 | (1.00-1.61) | 1.61 | (1.12-2.31) |
| Steroid tablets | 0.84 | (0.41-1.71) | .. | .. |
| Oestrogen only hormone replacement therapy | 1.28 | (0.61-2.70) | .. | .. |
| Antidepressants | 1.26 | (1.06-1.51) | 1.30 | (0.96-1.76) |
| Antipsychotics | 1.11 | (0.91-1.34) | 1.27 | (0.93-1.75) |
| Poor mobility/Parkinsons | 0.76 | (0.61-0.94) | 0.83 | (0.58-1.18) |
| Hearing impairment | 0.92 | (0.74-1.14) | 0.97 | (0.68-1.37) |
| Self-injurious behaviour | 0.64 | (0.36-1.15) | .. | .. |
| Visual impairment | 1.19 | (0.91-1.56) | 0.89 | (0.54-1.49) |
| Hypogonadism (including taking progesterone) | 1.42 | (1.03-1.95) | .. | .. |
| Sedatives and hypnotics | 0.86 | (0.68-1.08) | 1.03 | (0.71-1.48) |
| Proton pump inhibitors | 1.24 | (0.99-1.56) | 1.13 | (0.77-1.67) |
| Risperidone and/or hyperprolactinaemia | 1.06 | (0.76-1.47) | 1.01 | (0.58-1.76) |
| Type of intellectual disability<sup>b</sup> | .. | .. | .. | .. |
| Down syndrome | 1.03 | (0.79-1.35) | 1.93 | (1.28-2.91) |
| Other specified intellectual disability | 0.77 | (0.56-1.07) | 0.68 | (0.36-1.29) |
| Mild severity of intellectual disability<sup>c</sup> | 1.01 | (0.73-1.39) | 1.28 | (0.76-2.16) |

a Moderate/heavy drinking is defined as 3+ alcohol units daily
b The remainder of the cohort had an unspecified intellectual disability.
c The remainder of the cohort had non-mild or unspecified severity of intellectual disability.

Table 3 contains statistics from the internal validation. A comparison between apparent and optimism-corrected statistics indicated a small degree of optimism in both models. The optimism-corrected calibration slope was 0.966 (95% CI: 0.913-1.026) for the MOF model and 0.954 (95%CI: 0.888-1.027) for the HF model. These values were applied as uniform shrinkage coefficients to obtain the final prediction models (Table S3, Table S4).

**Table 3.** Validation statistics (95% confidence intervals) for the prediction models.

|  | **Apparent<sup>a</sup>** |  | **Optimism corrected<sup>b</sup>** |  | **External<sup>c</sup>** |  |
| --- | --- | --- | --- | --- | --- | --- |
| Major osteoporotic fracture model |  | .. | .. | .. | .. | .. |
| Calibration slope | 1.000 | .. | 0.966 | (0.913-1.026) | 0.982 | (0.947-1.020) |
| Harrell's C | 0.750 | (0.736-0.769) | 0.742 | (0.729-0.761) | 0.775 | (0.764-0.787) |
| Royston and Sauerbrei's $R^2_D$ | 0.388 | (0.362-0.431) | 0.368 | (0.343-0.411) | 0.428 | (0.407-0.449) |
| D discrimination | 1.628 | (1.543-1.780) | 1.560 | (1.475-1.712) | 1.771 | (1.697-1.848) |
| Hip fracture model |  |  |  |  |  |  |
| Calibration slope | 1.000 | .. | 0.954 | (0.888-1.027) | 0.968 | (0.928-1.012) |
| Harrell's C | 0.831 | (0.813-0.856) | 0.822 | (0.803-0.847) | 0.839 | (0.825-0.850) |
| Royston and Sauerbrei's $R^2_D$ | 0.542 | (0.513-0.593) | 0.518 | (0.488-0.569) | 0.542 | (0.519-0.564) |
| D discrimination | 2.228 | (2.100-2.472) | 2.117 | (1.988-2.360) | 2.225 | (2.125-2.329) |

a Apparent validation statistics show the performance of the model on the data from which it was derived.
b Apparent validation statistics were corrected for overfitting by subtracting an estimate of optimism. Optimism is the difference in performance on a bootstrap sample compared with the original sample for a model derived on the bootstrapped sample. The average optimism was calculated over 1000 bootstrap samples.
c External validation describes the performance of the final model on the Aurum cohort.

Statistics for the external validation (Table 3) and calibration plots (Figures 1 and 2) show that generally, both the HF and MOF models performed as well in the external validation data as in the derivation data. Calibration was good for both models, although there was a tendency for the predicted risk to be lower than the observed risk in some groups, representing the highest values of the predicted risk (Figures 1 and 2), and for the risk to be underestimated at intermediate values of the predicted risk. Harrell’s C and D statistics were 0.839 (95%CI: 0.825-0.850) and 2.225 (95%CI: 2.125-2.329) for the HF model. Statistics for the MOF model indicated that discrimination was not quite so good: Harrell’s C was 0.775 95%CI (0.764-0.787) and the D statistic was 1.771 (95% CI: 1.697-1.848). Calibration plots and Harrell’s C did not suggest any large degree of heterogeneity in IDFracture performance between subgroups (Figures S3-S10), although there were very few fractures in the youngest age group (aged 30-39 years) examined.

**Figure 1.**
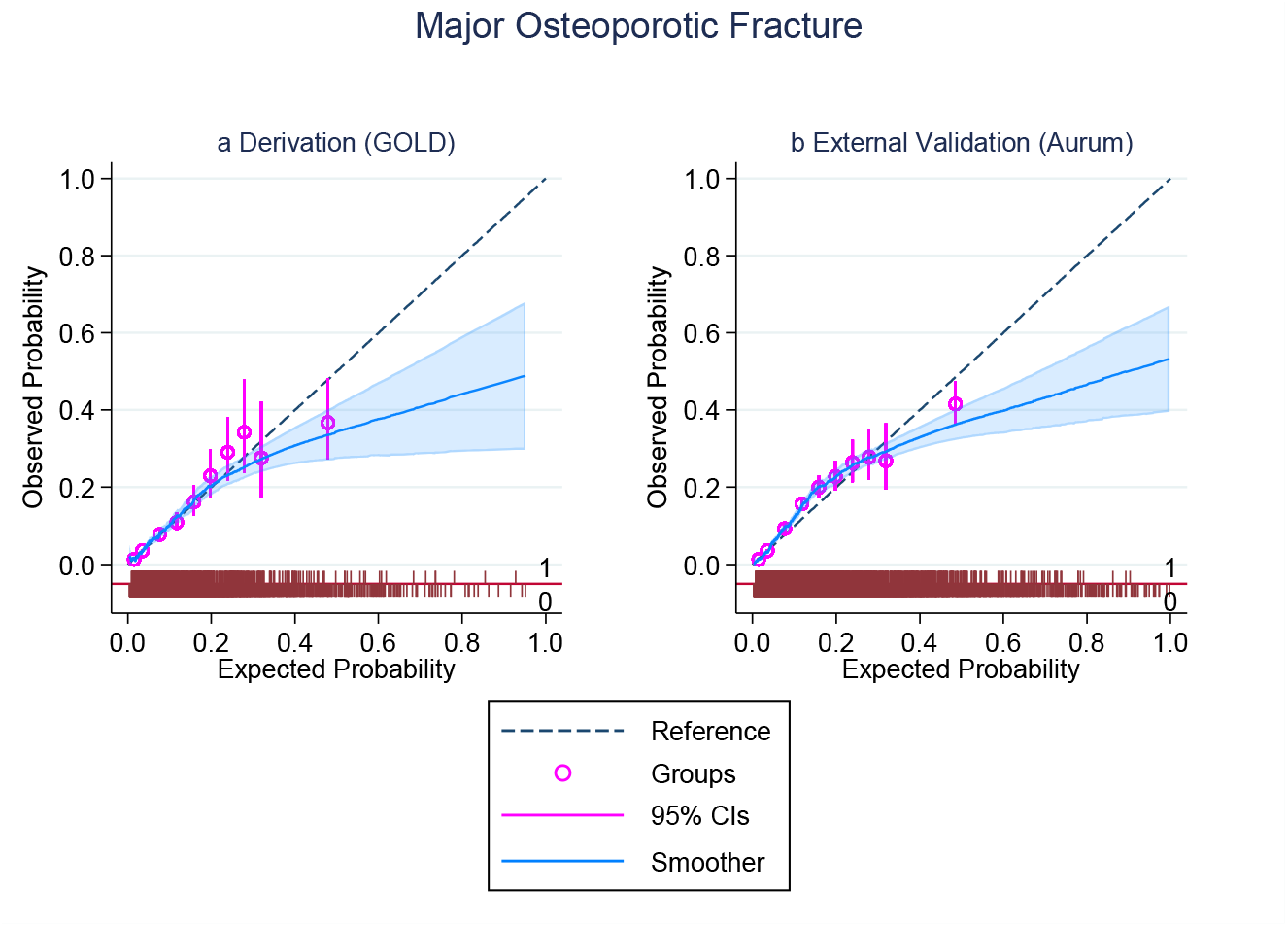
Calibration plot and distribution of 10-year predicted risks for major osteoporotic fracture (a) in the derivation cohort and (b) in the validation cohort. The observed 10-year risks are plotted against the predicted risks for each group of predicted risk (circles) and 95% confidence intervals (vertical lines). Ten groups were defined with cut-off points at predicted risks of 0.02, 0.06, 0.10, 0.14, 0.18, 0.22, 0.26, 0.30, and 0.34. The blue solid line and shaded area are the smoothed calibration curve and confidence interval, respectively. The dotted line indicates the equality of the observed and predicted risks. The spikes indicate the density of individuals who did (1) or did not (0) experience the outcome.

**Figure 2.**
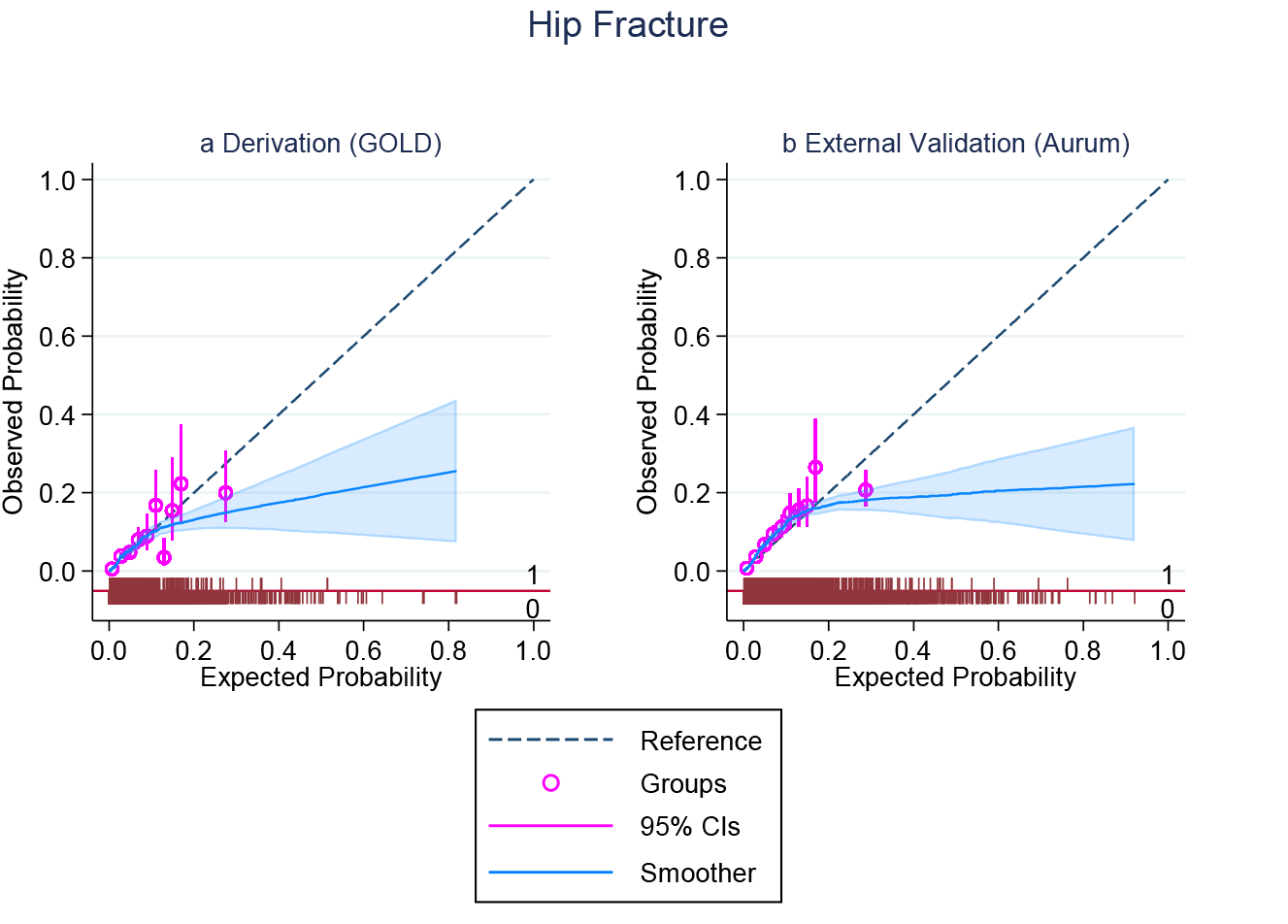
Calibration plot and distribution of 10-year predicted risks for hip fracture (a) in the derivation cohort and (b) in the validation cohort. The observed 10-year risks are plotted against the predicted risks for each group of predicted risk (circles) and 95% confidence intervals (vertical lines). Ten groups were defined with cut-off points at predicted risks of 0.02, 0.04, 0.06, 0.08, 0.10, 0.12, 0.14, 0.16, and 0.18. The blue solid line and shaded area are the smoothed calibration curve and confidence interval, respectively. The dotted line indicates the equality of the observed and predicted risks. The spikes indicate the density of individuals who did (1) or did not (0) experience the outcome.

## Discussion

We developed new prediction models (IDFracture) to predict the risk of MOF or HF in people with ID. To our knowledge, this is the first study to attempt to develop a fracture prediction model specifically for people with ID. We externally validated IDFracture via a more recent cohort derived from a different primary care database. All the models performed well in terms of both calibration and discrimination. The performance was also good in the sex, index year and IMD subgroups. It was less effective in some age groups.

The magnitudes of many of the HRs for predictors in our multivariate model were comparable to those found in the QFracture model [6]. Where different, they were accompanied by fairly wide confidence intervals. As with the QFracture model, epilepsy is a strong predictor for HFs or MOFs. However, we found that more than 20% of the ID population had a diagnosis of epilepsy or were on anticonvulsants, whereas epilepsy is a relatively uncommon condition in the general population. Epilepsy in people with ID is also very often treatment resistant and is also often associated with physical disabilities [24, 25]. We modelled the sex-specific relationships of age and fracture specifically for the ID population, as these associations are likely to differ from those in the general population. Among the additional predictors that we included, having Down syndrome, which occurred in 10.1% of the derivation cohort and 7.9% of the validation cohort, was a strong predictor of future HF, although it did not appear to be a strong predictor of MOF. People with Down syndrome have been repeatedly shown to have low bone mineral density, including in comparison with other people with ID, as well as deficits in bone formation [12, 26-28]. Down syndrome may also be associated with clinical characteristics such as hypogonadism, premature menopause, and coeliac disease, which are also risk factors for osteoporosis [13, 29, 30].

The Aurum database was released after we developed the prediction model in the relatively small GOLD database. We deliberately chose a more recent time period for external validation to test the model in a population most similar to that in which the prediction model would be used. There were some notable differences between the derivation and validation populations. First, the proportion of the population with Down syndrome was lower than that in the derivation population even after people who had only service user codes were excluded. Second, the prevalences of comorbidities and outcomes were greater. The median age of the validation cohort was slightly higher, consistent with increasing life expectancy. The introduction of the learning disability register in 2006 may have also changed the case mix and improved the diagnosis and treatment of this population [31]. The completeness of recording in clinical databases may also have improved overall over time.

The strengths of our study are the use of a large cohort with IDs, validation in a different database and a different time period. We included fractures recorded in HES inpatient data as well as in primary care data, reducing the risk of underestimating fracture incidence. We included additional risk factors that are relevant to the ID population. Risk factors were extracted from routinely collected primary care data, as IDFracture is designed for use in this setting. The limitations are that despite this being one of the largest database studies of people with ID, it was small compared with database studies performed in the general population, and many of our HRs had very wide confidence intervals. Other risk factors, such as smoking or ethnicity, could not be considered at the same level of detail as in models performed on the general population. Additionally, some variables were not well characterized. In particular, only a small percentage had a cause of ID or severity of ID recorded in their primary care records. The ID population itself is very heterogeneous, and a one-size-fits-all approach for predicting fracture risk may not be appropriate. Certain syndromes, e.g., Rett, Prader Willi, Williams, and aminoaciduria, are associated with extremely high rates of fracture, and it may not be appropriate to use IDFracture in such people. Finally, we did not consider the competing risk of death. Models that do not take this into account can overestimate the risk of the event of interest [32]. This is likely to be a particularly important consideration in the elderly population and may be more important in this ID population given the shorter life expectancy.

## Conclusion

Current guidelines for assessing the risk of fragility fracture do not specifically consider people with ID [33]. In general, people with ID have poorer bone health than the general population, and risk assessment tools designed for the general population are likely to underestimate risk. People with ID may also have difficulty accessing health services because of other comorbidities or behavioural or communication difficulties, so osteoporosis may remain undiagnosed until there is a fracture. We constructed and validated risk prediction models for HFs and MOFs in people with ID that are suitable for use in primary care. IDFracture could be used as a tool to identify people most likely to benefit from bone health assessment. It could be used during an annual health check, for example, the one that is offered in the United Kingdom to everyone on the learning disabilities register. We have recently completed a cost-effectiveness study of risk assessment strategies for MOFs and HFs that use IDFracture [34]. Further research related to the high risk of HF in people with Down syndrome should also be conducted.

## Data Availability

This study is based on an anonymised dataset from the Clinical Practice Research Datalink. Access rules to CPRD prevent researchers from sharing CPRD datasets. Data can be obtained via application to the Clinical Practice Research Datalink (https://www.cprd.com).

## Acknowledgements

We first of all acknowledge the leadership and vision of Dr Valeria Frighi, the grant holder, who died tragically in March 2024, following completion of the main work presented here. Without her inspiration and tireless work this study would never have happened. We also acknowledge the main charities relevant to this research, namely the Downs Syndrome Association, the Royal Osteoporosis Society, and Mencap, who have been extremely supportive of this work in recent years.

This project is funded by the National Institute for Health and Care Research (NIHR) under its Research for Patient Benefit (RfPB) Programme (Grant Reference Number PB-PG-1216-20017) and under its Policy Research (TRiP) Programme (Grant Reference Number NIHR202094). The funders of the study had no role in the study design, data collection, data analysis, data interpretation, or writing of the report. Professor Gary Collins is a National Institute for Health and Care Research (NIHR) Senior Investigator. The views expressed are those of the authors and not necessarily those of the NIHR or the Department of Health and Social Care.

This study is based in part on data from the Clinical Practice Research Datalink obtained under licence from the UK Medicines and Healthcare Products Regulatory Agency. The data are provided by patients and collected by the NHS as part of their care and support. The interpretation and conclusions contained in this study are those of the author/s alone. HES Data copyright © (2020), re-used with the permission of The Health & Social Care Information Centre. All rights reserved.

## Ethical approval

The study obtained ethics approval through the CPRD Research Data Governance process (Protocol Numbers 18_186 and 21_000433).

## References

1. Balogh R, Wood J, Dobranowski K, Lin E, Wilton A, Jaglal SB, Gemmill M, Lunsky Y (2017) Low-trauma fractures and bone mineral density testing in adults with and without intellectual and developmental disabilities: a population study. Osteoporos Int 28:727–732. 10.1007/s00198-016-3740-2

2. Büchele G, Becker C, Cameron ID, Auer R, Rothenbacher D, König HH, Rapp K (2017) Fracture risk in people with developmental disabilities: results of a large claims data analysis. Osteoporos Int 28:369–375. 10.1007/s00198-016-3733-1

3. Frighi V, Smith M, Andrews TM, Clifton L, Collins GS, Fuller A, Roast J, Holt TA (2022) Incidence of fractures in people with intellectual disabilities over the life course: a retrospective matched cohort study. eClinicalMedicine 52:101656. 10.1016/j.eclinm.2022.101656

4. Whitney DG, Caird MS, Jepsen KJ, Kamdar NS, Marsack-Topolewski CN, Hurvitz EA, Peterson MD (2020) Elevated fracture risk for adults with neurodevelopmental disabilities. Bone 130:115080. 10.1016/j.bone.2019.115080

5. Kanis JA, Johnell O, Oden A, Johansson H, McCloskey E (2008) FRAX™ and the assessment of fracture probability in men and women from the UK. Osteoporos Int 19:385–397. 10.1007/s00198-007-0543-5

6. Hippisley-Cox J, Coupland C (2012) Derivation and validation of updated QFracture algorithm to predict risk of osteoporotic fracture in primary care in the United Kingdom: prospective open cohort study. Br Med J 344:e3427. 10.1136/bmj.e3427

7. Herrett E, Gallagher AM, Bhaskaran K, Forbes H, Mathur R, van Staa T, Smeeth L (2015) Data Resource Profile: Clinical Practice Research Datalink (CPRD). Int J Epidemiol 44:827–836. 10.1093/ije/dyv098

8. Wolf A, Dedman D, Campbell J, Booth H, Lunn D, Chapman J, Myles P (2019) Data resource profile: Clinical Practice Research Datalink (CPRD) Aurum. Int J Epidemiol 48:1740–1740g. 10.1093/ije/dyz034

9. Collins GS, Moons KGM, Dhiman P, et al. (2024) TRIPOD+AI statement: updated guidance for reporting clinical prediction models that use regression or machine learning methods. Br Med J 385:e078378. 10.1136/bmj-2023-078378

10. StataCorp (2019) Stata 16. College Station, TX: StataCorp LLC

11. Curtis EM, van der Velde R, Moon RJ, van den Bergh JPW, Geusens P, de Vries F, van Staa TP, Cooper C, Harvey NC (2016) Epidemiology of fractures in the United Kingdom 1988–2012: Variation with age, sex, geography, ethnicity and socioeconomic status. Bone 87:19–26. 10.1016/j.bone.2016.03.006

12. Hawli Y, Nasrallah M, Fuleihan GE-H (2009) Endocrine and musculoskeletal abnormalities in patients with Down syndrome. Nat Rev Endocrinol 5:327–334. 10.1038/nrendo.2009.80

13. Schupf N, Zigman W, Kapell D, Lee JH, Kline J, Levin B (1997) Early menopause in women with Down’s syndrome. J Intellect Disabil Res 41:264–267. 10.1046/j.1365-2788.1997.03838.x

14. Frighi V, Stephenson MT, Morovat A, et al. (2011) Safety of antipsychotics in people with intellectual disability. Br J Psychiatry 199:289–295. 10.1192/bjp.bp.110.085670

15. Arvio M, Kilpinen-Loisa P, Tiitinen A, Huovinen K, MÄKitie O (2009) Bone mineral density and sex hormone status in intellectually disabled women on progestin-induced amenorrhea. Acta Obstet Gynecol Scand 88:428–433. 10.1080/00016340902763244

16. Frighi V, Morovat A, Andrews TM, Rana F, Stephenson MT, White SJ, Fower E, Roast J, Goodwin GM (2019) Vitamin D, bone mineral density and risk of fracture in people with intellectual disabilities. J Intellect Disabil Res 63:357–367. 10.1111/jir.12581

17. Holick MF (2007) Vitamin D deficiency. N Engl J Med 357:266–281. 10.1056/NEJMra070553

18. Morris TR, Patrick (2015) MFPMI: Stata module to build multivariable fractional polynomial models in multiply imputed data. revised 15 Feb 2016 edn. Statistical Software Components S458021, Boston College Department of Economics,

19. Steyerberg EW (2019) Clinical Prediction Models A Practical Approach to Development, Validation, and Updating. Springer Cham. 10.1007/978-3-030-16399-0

20. Ensor J. Kis, E.C. Martin (2018) “PMCALPLOT: Stata module to produce calibration plot of prediction model performance,” Statistical Software Components S458486, Boston College Department of Economics. revised 08 Mar 2024 edn

21. Royston P (2006) Explained Variation for Survival Models. The Stata Journal 6:83–96. 10.1177/1536867X0600600105

22. Royston P, Sauerbrei W (2004) A new measure of prognostic separation in survival data. Stat Med 23:723–748. 10.1002/sim.1621

23. Noma H, Shinozaki T, Iba K, Teramukai S, Furukawa TA (2021) Confidence intervals of prediction accuracy measures for multivariable prediction models based on the bootstrap-based optimism correction methods. Stat Med 40:5691–5701. 10.1002/sim.9148

24. Robertson J, Hatton C, Emerson E, Baines S (2015) Prevalence of epilepsy among people with intellectual disabilities: A systematic review. Seizure 29:46–62. 10.1016/j.seizure.2015.03.016

25. McGrother CW, Bhaumik S, Thorp CF, Hauck A, Branford D, Watson JM (2006) Epilepsy in adults with intellectual disabilities: Prevalence, associations and service implications. Seizure 15:376–386. 10.1016/j.seizure.2006.04.002

26. Geijer JR, Stanish HI, Draheim CC, Dengel DR (2014) Bone mineral density in adults with Down syndrome, intellectual disability, and nondisabled adults. Am J Intellect Dev Disabil 119:107–114. 10.1352/1944-7558-119.2.107

27. Carfì A, Liperoti R, Fusco D, et al. (2017) Bone mineral density in adults with Down syndrome. Osteoporos Int 28:2929–2934. 10.1007/s00198-017-4133-x

28. LaCombe JM, Roper RJ (2020) Skeletal dynamics of Down syndrome: A developing perspective. Bone 133:115215. 10.1016/j.bone.2019.115215

29. Ejskjaer K, Uldbjerg N, Goldstein H (2006) Menstrual profile and early menopause in women with Down syndrome aged 26–40 years. J Intellect Dev Disabil 31:166–171. 10.1080/13668250600879222

30. Du Y, Shan LF, Cao ZZ, Feng JC, Cheng Y (2018) Prevalence of celiac disease in patients with Down syndrome: a meta-analysis. Oncotarget 9:5387–5396. 10.18632/oncotarget.23624

31. Hatton GG, Emerson E., Brown I. (2016) Learning Disabilities Observatory. People with learning disabilities in England 2015: Main report. PHE publications gateway number: 2016404. In England Ph (ed)

32. Wolbers M, Koller MT, Witteman JC, Steyerberg EW (2009) Prognostic models with competing risks: methods and application to coronary risk prediction. Epidemiol 20:555–561. 10.1097/EDE.0b013e3181a39056

33. NICE (2012) Osteoporosis: assessing the risk of fragility fracture. https://www.nice.org.uk/guidance/cg146 xAccessed 27 June 2024

34. Png ME, Frighi V, Holt TA, Achana F, Smith M, Collins GS, Roast J, Petrou S. Cost-effectiveness of osteoporotic fracture risk assessment in people with intellectual disabilities. Submitted to Osteoporosis International.

