## Supplementary material for "Development and external validation of prediction models for major osteoporotic fracture and hip fracture in people with intellectual disability"

Table S1 Univariate and age and gender adjusted hazard ratios for predictors of hip fracture

Table S2 Univariate and age and gender adjusted hazard ratios for predictors of major osteoporotic fracture (MOF)

Table S3 ID fracture prediction models

Figure S1 Kaplan Meier curves for cumulative incidence of hip fracture in the GOLD derivation cohort and in the Aurum validation cohort

Figure S2 Kaplan Meier curves for cumulative incidence of major osteoporotic fracture (MOF) in the GOLD derivation cohort and in the Aurum validation cohort

Figure S3 Calibration plots for the performance of the major osteoporotic fracture (MOF) prediction model in the Aurum validation data. Groups are deciles of predicted risk. Validation was done by subgroup of age at index.

Figure S4 Calibration plots for the performance of the major osteoporotic fracture (MOF) prediction model in the Aurum validation data. Groups are deciles of predicted risk. Validation was done by subgroup of sex.

Figure S5 Calibration plots for the performance of the major osteoporotic fracture (MOF) prediction model in the Aurum validation data. Groups are deciles of predicted risk. Validation was done by subgroup of Index of Multiple Deprivation.

Figure S6 Calibration plots for the performance of the major osteoporotic fracture (MOF) prediction model in the Aurum validation data. Groups are deciles of predicted risk. Validation was done by subgroup of year of index date.

Figure S7 Calibration plots for the performance of the hip fracture prediction model in the Aurum validation data. Groups are deciles of predicted risk. Validation was done by subgroup of age at index.

Figure S8 Calibration plots for the performance of the hip fracture prediction model in the Aurum validation data. Groups are deciles of predicted risk. Validation was done by subgroup of sex.

Figure S9 Calibration plots for the performance of the hip fracture prediction model in the Aurum validation data. Groups are deciles of predicted risk. Validation was done by subgroup of Index of Multiple Deprivation.

Figure S10 Calibration plots for the performance of the hip fracture prediction model in the Aurum validation data. Groups are deciles of predicted risk. Validation was done by subgroup of year of index date.

**Table S1. Hazard ratios (95% confidence intervals) for predictors of hip fracture in the derivation (360 hip fractures) and validation cohorts (1001 hip fractures). Hazard ratios were estimated using univariate Cox regression models or in models containing age and gender in addition to the predictor variable.**

| Predictor | Crude | | | | Adjusted for age and gender | | | |
|---|---|---|---|---|---|---|---|---|
| | Derivation cohort | | Validation cohort | | Derivation cohort | | Validation cohort | |
| | HR | (95% CI) | HR | (95% CI) | HR | (95% CI) | HR | (95% CI) |
| Ten years higher age | 2·23 | (2·07-2·40) | 2·31 | (2·20-2·42) | 2·22 | (2·06-2·39) | 2·31 | (2·20-2·42) |
| Female | 1·33 | (1·08-1·63) | 1·22 | (1·07-1·38) | 1·15 | (0·93-1·41) | 1·05 | (0·92-1·18) |
| Five units higher BMI ($kg/m^2$) | 0·76 | (0·67-0·85) | 0·71 | (0·67-0·76) | 0·73 | (0·65-0·83) | 0·72 | (0·67-0·76) |
| Ten cm taller | 0·83 | (0·74-0·93) | 0·81 | (0·77-0·86) | 0·93 | (0·80-1·09) | 0·90 | (0·83-0·96) |
| Current smoker | 0·78 | (0·58-1·07) | 0·89 | (0·75-1·05) | 0·85 | (0·62-1·15) | 1·05 | (0·89-1·24) |
| Current moderate/ heavy drinker[a] | 0·90 | (0·62-1·30) | 0·73 | (0·49-1·09) | 1·06 | (0·73-1·54) | 0·93 | (0·62-1·38) |
| Previous major osteoporotic fracture (MOF) | 2·91 | (2·15-3·94) | 4·53 | (3·85-5·34) | 2·09 | (1·54-2·84) | 2·58 | (2·18-3·05) |
| Previous fracture at non-MOF site | 1·31 | (0·99-1·73) | 1·38 | (1·19-1·58) | 1·40 | (1·06-1·84) | 1·43 | (1·24-1·64) |
| Dementia | 8·01 | (4·38-14·62) | 4·78 | (3·53-6·48) | 2·19 | (1·19-4·02) | 1·58 | (1·16-2·15) |
| History of falls | 3·42 | (2·51-4·66) | 3·56 | (3·06-4·15) | 2·17 | (1·58-2·97) | 2·04 | (1·75-2·39) |
| Asthma or chronic obstructive pulmonary disease | 0·95 | (0·66-1·36) | 0·78 | (0·64-0·96) | 1·07 | (0·75-1·55) | 0·92 | (0·75-1·12) |
| Cancer | 4·13 | (2·32-7·34) | 2·38 | (1·76-3·22) | 1·86 | (1·04-3·33) | 1·08 | (0·79-1·46) |
| Heart disease | 2·17 | (1·25-3·78) | 2·88 | (2·27-3·64) | 0·70 | (0·40-1·23) | 1·09 | (0·85-1·38) |
| Epilepsy or taking anticonvulsants | 1·65 | (1·32-2·07) | 1·63 | (1·43-1·85) | 2·00 | (1·59-2·51) | 1·68 | (1·48-1·91) |
| Liver or kidney disease | 1·83 | (0·86-3·86) | 2·71 | (2·08-3·53) | 1·14 | (0·54-2·40) | 1·02 | (0·78-1·33) |
| Diabetes | 2·64 | (1·86-3·74) | 2·03 | (1·69-2·44) | 1·42 | (1·00-2·02) | 1·11 | (0·92-1·33) |
| Antidepressants | 1·60 | (1·21-2·11) | 1·25 | (1·08-1·46) | 1·37 | (1·04-1·81) | 1·07 | (0·92-1·25) |
| Antipsychotics | 1·59 | (1·21-2·08) | 1·75 | (1·52-2·01) | 1·31 | (1·00-1·72) | 1·42 | (1·23-1·64) |
| Poor mobility/Parkinsons | 1·11 | (0·79-1·57) | 1·54 | (1·31-1·81) | 1·16 | (0·82-1·64) | 1·37 | (1·17-1·61) |
| Hearing impairment | 1·16 | (0·82-1·64) | 1·19 | (1·00-1·42) | 1·07 | (0·76-1·51) | 1·06 | (0·89-1·26) |
| Visual impairment | 1·19 | (0·72-1·96) | 1·33 | (1·08-1·64) | 1·03 | (0·62-1·69) | 1·10 | (0·90-1·36) |
| Sedatives and hypnotics | 1·55 | (1·09-2·19) | 1·89 | (1·57-2·27) | 1·30 | (0·92-1·85) | 1·59 | (1·32-1·91) |
| Proton pump inhibitors | 1·98 | (1·37-2·86) | 2·16 | (1·85-2·53) | 1·28 | (0·88-1·85) | 1·31 | (1·12-1·54) |
| Risperidone and/or hyperprolactinaemia | 1·28 | (0·78-2·08) | 1·37 | (1·10-1·71) | 1·30 | (0·80-2·12) | 1·30 | (1·04-1·62) |
| Type of intellectual disability[b] | .. | .. | .. | .. | .. | .. | .. | .. |
| Down syndrome | 1·00 | (0·71-1·40) | 1·16 | (0·94-1·44) | 1·58 | (1·12-2·23) | 1·65 | (1·33-2·05) |
| Other specified intellectual disability | 0·52 | (0·27-0·97) | 0·77 | (0·59-1·02) | 0·62 | (0·33-1·17) | 0·96 | (0·73-1·28) |
| Mild severity of intellectual disability[c] | 1·27 | (0·76-2·13) | 0·79 | (0·64-0·97) | 1·17 | (0·70-1·96) | 0·84 | (0·68-1·03) |

a Moderate/heavy drinking is defined as 3+ alcohol units daily

b The remainder of the cohort had an unspecified intellectual disability.

c The remainder of the cohort had non-mild or unspecified severity of intellectual disability.

**Table S2. Hazard ratios (HR) (95% confidence intervals (CI)) for predictors of major osteoporotic fracture (MOF) in the derivation (1045 MOF) and validation cohorts (2420 MOF). Hazard ratios were estimated using univariate Cox regression models or with models containing age and gender in addition to the predictor variable.**

| Predictor | Unadjusted | | | | Adjusted for age and gender | | | |
|---|---|---|---|---|---|---|---|---|
| | Derivation cohort | | Validation cohort | | Derivation cohort | | Validation cohort | |
| | HR | (95% CI) | HR | (95% CI) | HR | (95% CI) | HR | (95% CI) |
| Ten years older age | 1.61 | (1.42-1.82) | 1.86 | (1.81-1.92) | 1.48 | (1.31-1.67) | 1.84 | (1.79-1.90) |
| Female at age 40 years | 1.74 | (1.66-1.81) | 1.56 | (1.44-1.69) | 1.72 | (1.65-1.79) | 1.41 | (1.30-1.53) |
| Five units higher body mass index (kg/m2) | 0.91 | (0.86-0.97) | 0.85 | (0.82-0.88) | 0.88 | (0.82-0.94) | 0.84 | (0.81-0.87) |
| 10cm taller | 0.85 | (0.79-0.91) | 0.83 | (0.80-0.86) | 1.01 | (0.91-1.11) | 0.99 | (0.94-1.04) |
| Current smoker | 0.91 | (0.77-1.08) | 0.94 | (0.84-1.04) | 0.98 | (0.83-1.17) | 1.09 | (0.98-1.22) |
| Current moderate/ heavy drinker[a] | 1.17 | (0.97-1.43) | 1.12 | (0.91-1.38) | 1.33 | (1.09-1.61) | 1.46 | (1.19-1.81) |
| Non-white ethnicity | 0.44 | (0.24-0.79) | 0.39 | (0.32-0.48) | 0.60 | (0.33-1.09) | 0.54 | (0.44-0.66) |
| Previous major osteoporotic fracture (MOF) | 3.51 | (2.97-4.15) | 4.47 | (4.01-4.97) | 2.87 | (2.42-3.40) | 2.92 | (2.62-3.26) |
| Previous fracture at non-MOF site | 1.43 | (1.22-1.68) | 1.50 | (1.37-1.64) | 1.52 | (1.30-1.78) | 1.59 | (1.45-1.73) |
| Dementia | 4.79 | (3.07-7.46) | 3.66 | (2.93-4.57) | 1.84 | (1.17-2.87) | 1.46 | (1.17-1.84) |
| History of falls | 3.02 | (2.50-3.66) | 3.26 | (2.94-3.61) | 2.16 | (1.78-2.62) | 2.12 | (1.91-2.35) |
| Malabsorption | 1.40 | (0.78-2.54) | 1.34 | (1.01-1.79) | 1.45 | (0.80-2.63) | 1.31 | (0.98-1.75) |
| Endocrine problems | 2.22 | (1.37-3.58) | 1.61 | (1.19-2.18) | 1.57 | (0.97-2.54) | 1.20 | (0.89-1.64) |
| Asthma or chronic obstructive pulmonary disease | 1.19 | (0.98-1.44) | 1.08 | (0.96-1.21) | 1.28 | (1.05-1.55) | 1.22 | (1.09-1.36) |
| Cancer | 3.03 | (2.05-4.47) | 2.05 | (1.66-2.53) | 1.68 | (1.13-2.48) | 1.09 | (0.89-1.35) |
| Heart disease | 1.76 | (1.23-2.52) | 2.54 | (2.16-2.98) | 0.78 | (0.54-1.13) | 1.23 | (1.04-1.45) |
| Epilepsy or taking anticonvulsants | 1.62 | (1.42-1.85) | 1.77 | (1.63-1.92) | 1.82 | (1.59-2.08) | 1.78 | (1.64-1.93) |
| Rheumatoid arthritis or systemic lupus erythematosus | 2.74 | (1.47-5.11) | 2.98 | (2.07-4.30) | 1.66 | (0.89-3.09) | 1.94 | (1.35-2.80) |
| Liver disease | 1.46 | (0.81-2.64) | 3.39 | (1.92-5.98) | 1.36 | (0.75-2.46) | 2.68 | (1.52-4.72) |
| Chronic kidney disease | 3.06 | (1.73-5.42) | 2.08 | (1.71-2.54) | 1.42 | (0.80-2.51) | 0.91 | (0.74-1.11) |
| Diabetes | 2.01 | (1.60-2.53) | 1.71 | (1.51-1.95) | 1.27 | (1.01-1.60) | 1.06 | (0.93-1.20) |
| Steroid tablets | 1.41 | (0.70-2.83) | 2.58 | (1.90-3.50) | 0.99 | (0.49-1.98) | 1.92 | (1.41-2.60) |
| Oestrogen only hormone replacement therapy | 2.03 | (0.97-4.27) | 1.07 | (0.53-2.14) | 1.46 | (0.69-3.08) | 0.88 | (0.44-1.76) |
| Antidepressants | 1.58 | (1.34-1.86) | 1.46 | (1.33-1.61) | 1.36 | (1.15-1.60) | 1.25 | (1.13-1.37) |
| Antipsychotics | 1.38 | (1.17-1.63) | 1.46 | (1.33-1.60) | 1.19 | (1.01-1.41) | 1.22 | (1.11-1.34) |
| Poor mobility/Parkinsons | 1.02 | (0.83-1.25) | 1.37 | (1.23-1.52) | 1.04 | (0.85-1.28) | 1.25 | (1.12-1.39) |
| Hearing impairment | 1.06 | (0.86-1.30) | 1.17 | (1.04-1.31) | 1.01 | (0.82-1.24) | 1.09 | (0.97-1.22) |
| Self-injurious behaviour | 0.69 | (0.39-1.23) | 1.48 | (1.15-1.91) | 0.84 | (0.47-1.48) | 1.77 | (1.37-2.28) |
| Visual impairment | 1.46 | (1.12-1.91) | 1.36 | (1.19-1.55) | 1.34 | (1.02-1.75) | 1.17 | (1.03-1.34) |
| Hypogonadism (including taking progesterone) | 1.02 | (0.75-1.39) | 0.86 | (0.70-1.04) | 1.49 | (1.09-2.05) | 1.21 | (0.99-1.48) |
| Sedatives and hypnotics | 1.26 | (1.01-1.57) | 1.69 | (1.49-1.91) | 1.09 | (0.87-1.36) | 1.44 | (1.27-1.63) |
| Proton pump inhibitors | 1.92 | (1.54-2.38) | 2.05 | (1.85-2.27) | 1.41 | (1.14-1.76) | 1.39 | (1.25-1.55) |
| Risperidone and/or hyperprolactinaemia | 1.23 | (0.92-1.64) | 1.20 | (1.03-1.39) | 1.21 | (0.91-1.62) | 1.12 | (0.96-1.30) |
| Type of intellectual disability[b] | | | | | | | | |
| Down syndrome | 0.69 | (0.55-0.87) | 0.85 | (0.73-1.00) | 0.86 | (0.68-1.09) | 1.02 | (0.87-1.20) |
| Other specified intellectual disability | 0.68 | (0.49-0.94) | 0.88 | (0.74-1.04) | 0.76 | (0.55-1.06) | 1.03 | (0.87-1.22) |
| Mild severity of intellectual disability[c] | 1.10 | (0.80-1.53) | 0.93 | (0.82-1.05) | 1.06 | (0.77-1.46) | 0.96 | (0.85-1.09) |

a Moderate/heavy drinking is defined as 3+ alcohol units daily; b The remainder of the cohort had an unspecified intellectual disability.
c The remainder of the cohort had non-mild or unspecified severity of intellectual disability.

**Table S3. IDfracture prediction models for hip and major osteoporotic fracture (MOF)**

| | Major osteoporotic fracture | Hip fracture |
|---|---|---|
| Coefficents | ·· | ·· |
| (Age at index (years) - 40)/10 in females | 0.5604 | 0.8330 |
| (Age at index (years) - 40)/10 in males | 0.4438 | 0.7334 |
| Female | 0.2192 | -0.0561 |
| (Body mass index(kg/m2) - 25)/5 | -0.1505 | -0.3473 |
| (Height (cm) - 165)/10 | -0.0265 | -0.0399 |
| Current smoker | -0.0845 | -0.1783 |
| Current moderate/ heavy drinker[a] | 0.2473 | 0.0872 |
| Non-white ethnicity | -0.4247 | ·· |
| Previous major osteoporotic fracture (MOF) | 0.9947 | 0.6359 |
| Previous fracture at non-MOF site | 0.4712 | 0.3725 |
| Dementia | 0.2724 | 0.2640 |
| History of falls | 0.5000 | 0.5514 |
| Malabsorption | 0.3340 | ·· |
| Endocrine problems | 0.3431 | ·· |
| Asthma or chronic obstructive pulmonary disease | 0.1572 | 0.0584 |
| Cancer | 0.3982 | 0.5716 |
| Heart disease | -0.3720 | -0.4186 |
| Epilepsy or taking anticonvulsants | 0.5503 | 0.6383 |
| Rheumatoid arthritis or systemic lupus erythematosus | 0.4116 | ·· |
| Liver disease | 0.1401 | ·· |
| Chronic kidney disease | 0.2088 | ·· |
| Liver or kidney disease | | 0.0208 |
| Diabetes | 0.2382 | 0.4749 |
| Steroid tablets | -0.1751 | ·· |
| Oestrogen only hormone replacement therapy | 0.2464 | ·· |
| Antidepressants | 0.2349 | 0.2637 |
| Antipsychotics | 0.1003 | 0.2419 |
| Poor mobility/Parkinsons | -0.2750 | -0.1886 |
| Hearing impairment | -0.0844 | -0.0338 |
| Self-injurious behaviour | -0.4389 | ·· |
| Visual impairment | 0.1736 | -0.1111 |
| Hypogonadism (including taking progesterone) | 0.3480 | ·· |
| Sedatives and hypnotics | -0.1530 | 0.0256 |
| Proton pump inhibitors | 0.2140 | 0.1254 |
| Risperidone and/or hyperprolactinaemia | 0.0557 | 0.0114 |
| Down syndrome | 0.0296 | 0.6577 |
| Other specified intellectual disability[b] | -0.2616 | -0.3791 |
| Mild severity of intellectual disability[c] | 0.0063 | 0.2469 |
| Optimism correction | 0.9657 | 0.9539 |
| 10-year baseline survival | 0.9773 | 0.9934 |

The 10-year risk of fracture for an individual is calculated as $1 - [S_{0t}{}^{\exp(LP)}]$ where $S_{0t}$ is the 10-year baseline survival and LP is the linear predictor. LP is calculated by multiplying the value of each variable by its coefficient, and then summing over all variables. The value of binary variables is 1 if they have the condition, otherwise 0. Continuous variables must be scaled as described in the table. After summing the optimism corrected LP is calculated by multiplying by the optimism correction term.

a Moderate/heavy drinking is defined as 3+ alcohol units daily
b The remainder of the cohort had an unspecified intellectual disability.
c The remainder of the cohort had non-mild or unspecified severity of intellectual disability.

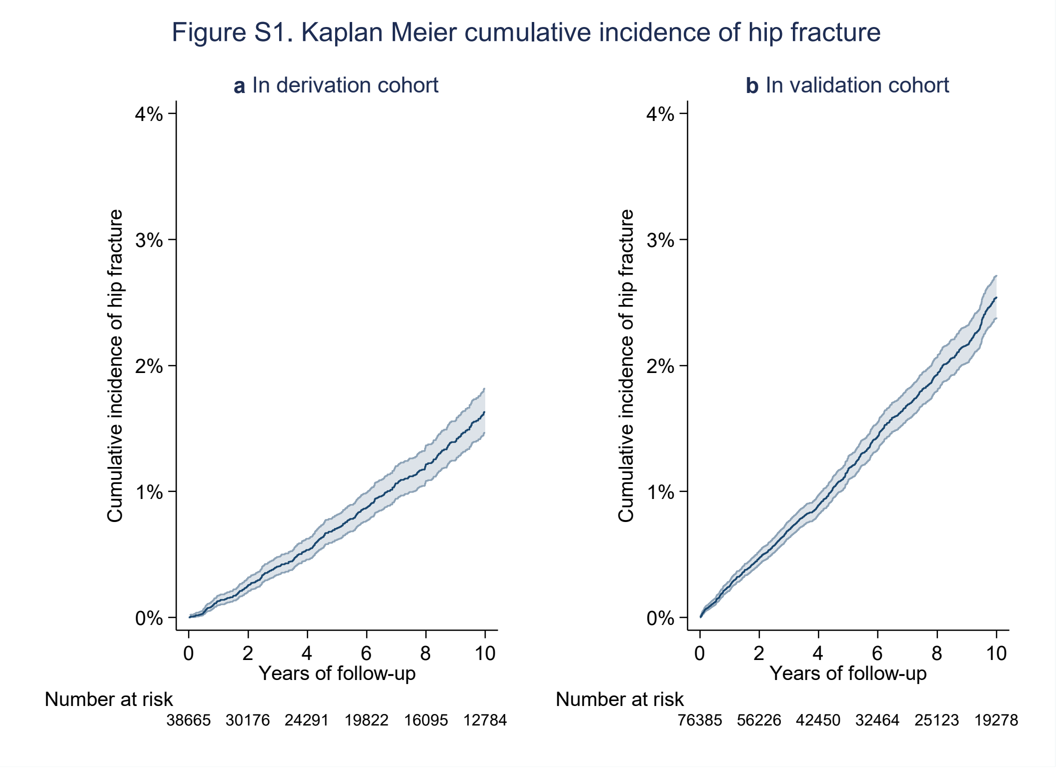


Figure S1 Kaplan Meier curves for cumulative incidence of hip fracture in the GOLD derivation cohort and in the Aurum validation cohort

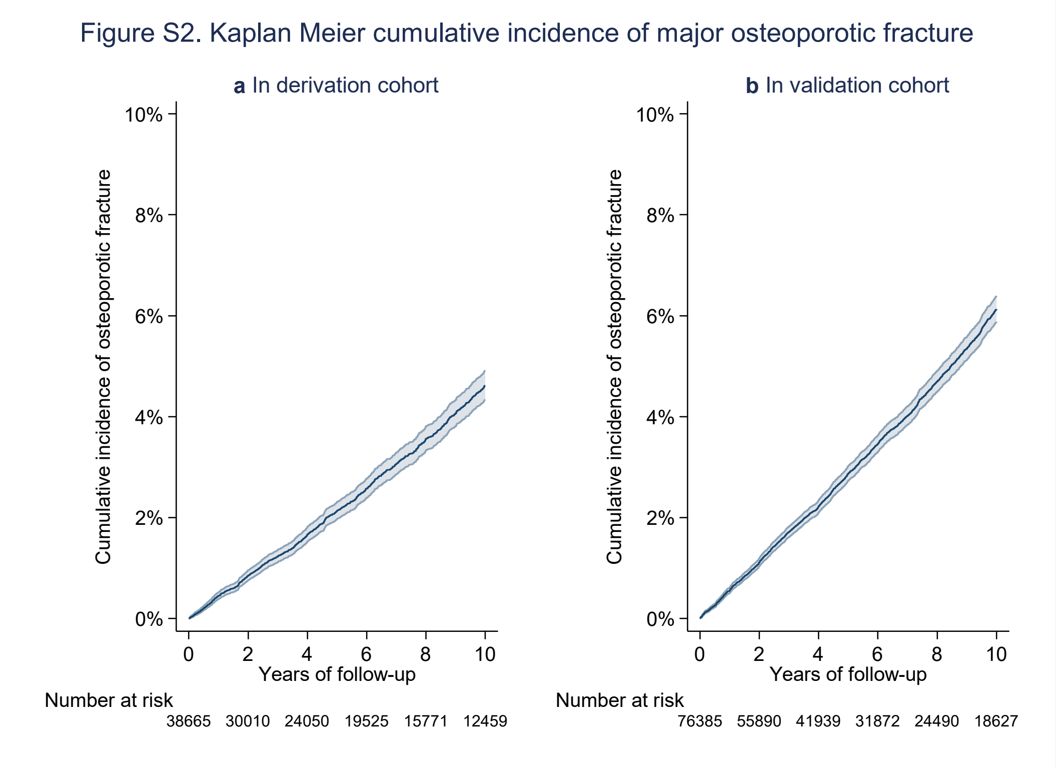


Figure S2 Kaplan Meier curves for cumulative incidence of major osteoporotic fracture (MOF) in the GOLD derivation cohort and in the Aurum validation cohort

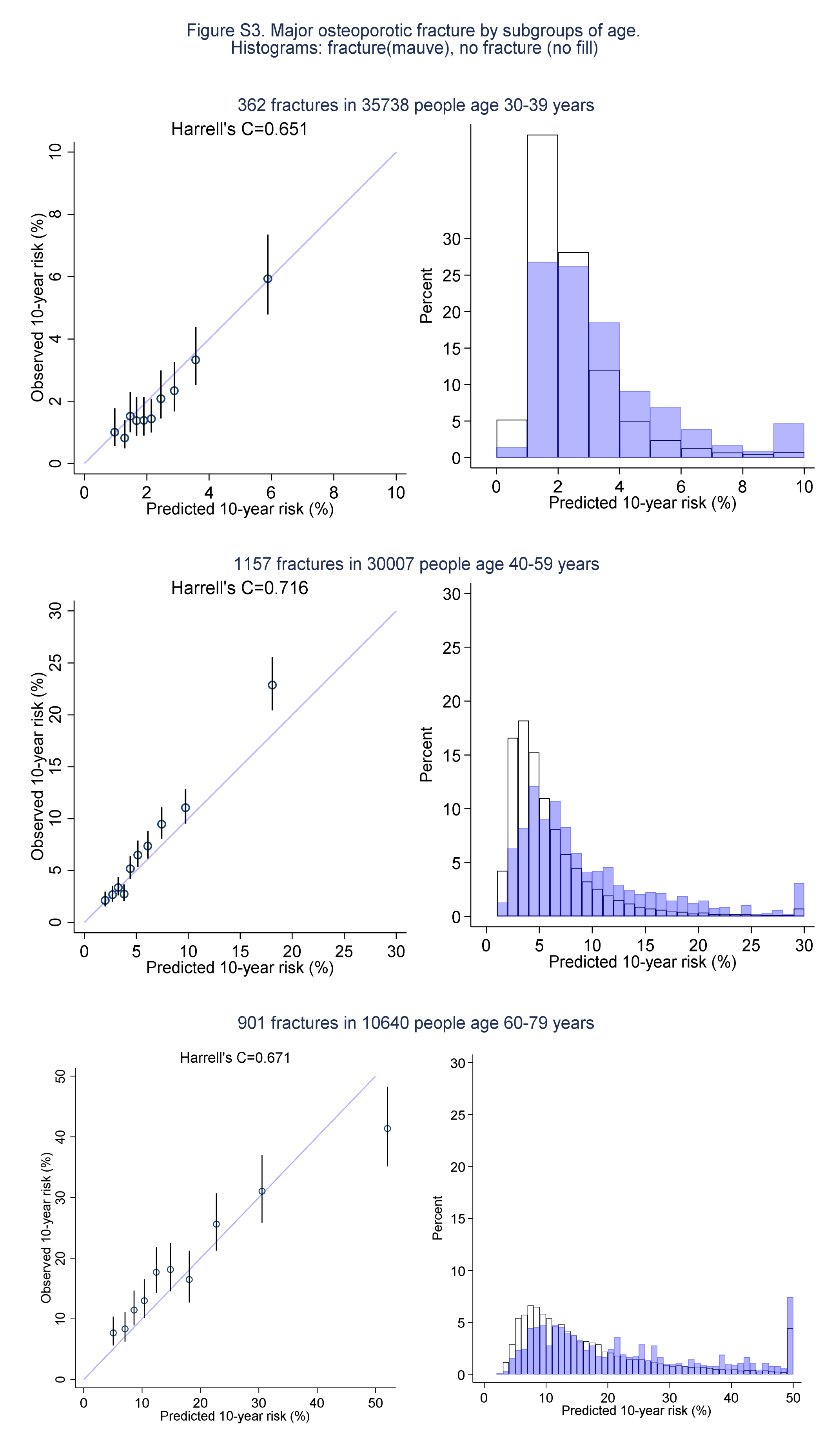


Figure S3 Calibration plots for the performance of the major osteoporotic fracture (MOF) prediction model in the Aurum validation data. Groups are deciles of predicted risk. Validation was done by subgroup of age at index.

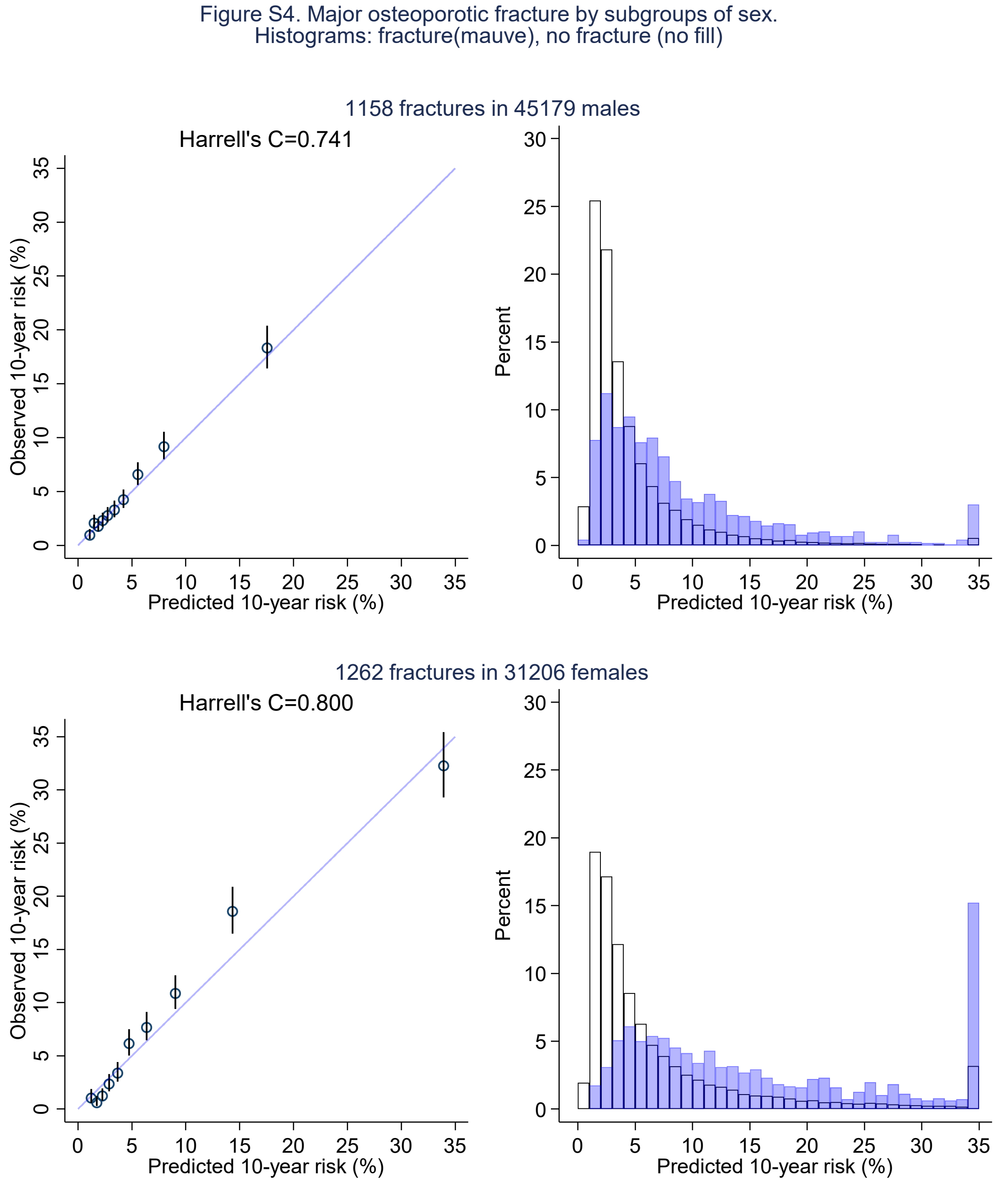


Figure S4 Calibration plots for the performance of the major osteoporotic fracture (MOF) prediction model in the Aurum validation data. Groups are deciles of predicted risk. Validation was done by subgroup of sex.

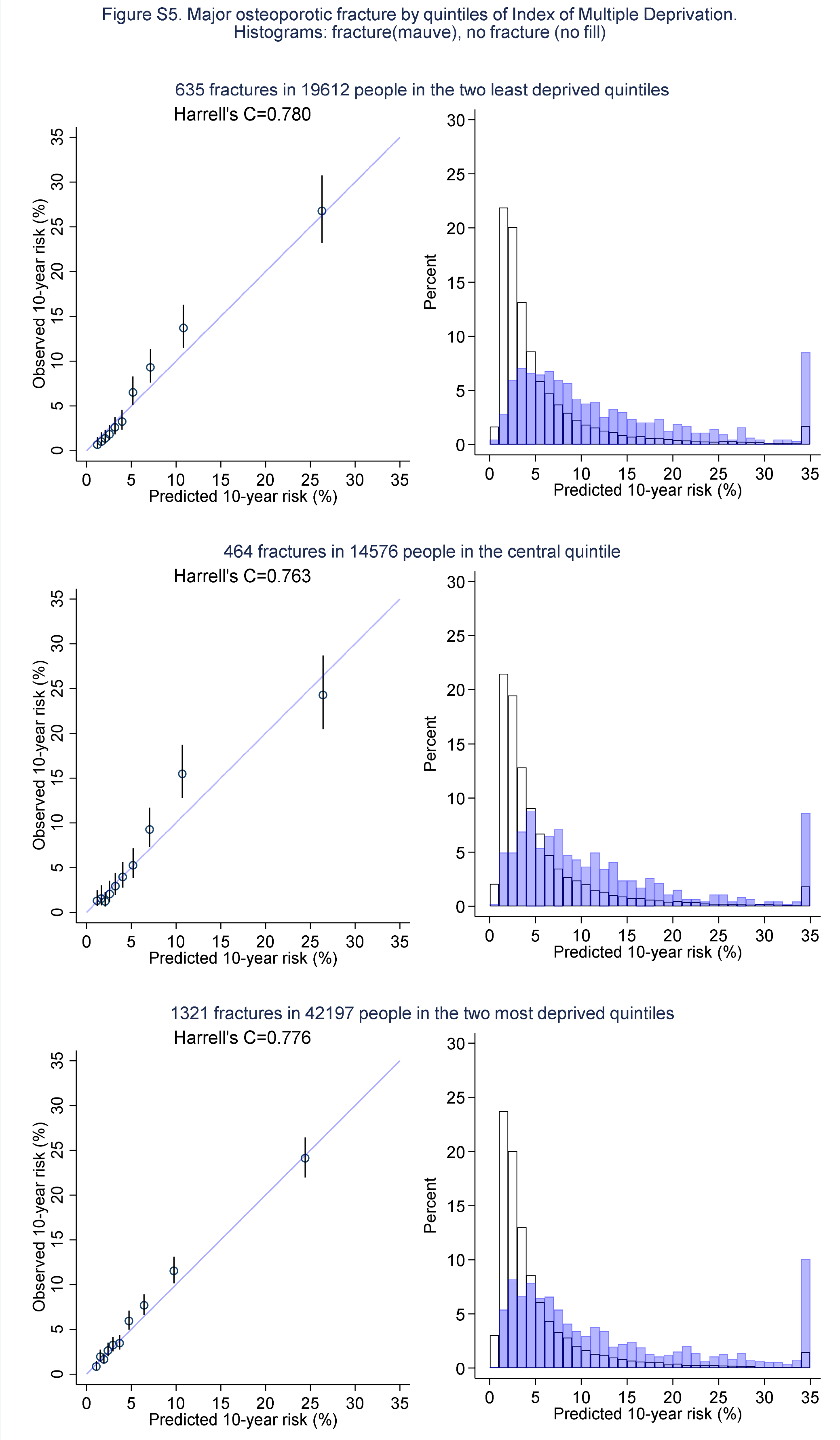


Figure S5 Calibration plots for the performance of the major osteoporotic fracture (MOF) prediction model in the Aurum validation data. Groups are deciles of predicted risk. Validation was done by subgroup of Index of Multiple Deprivation.

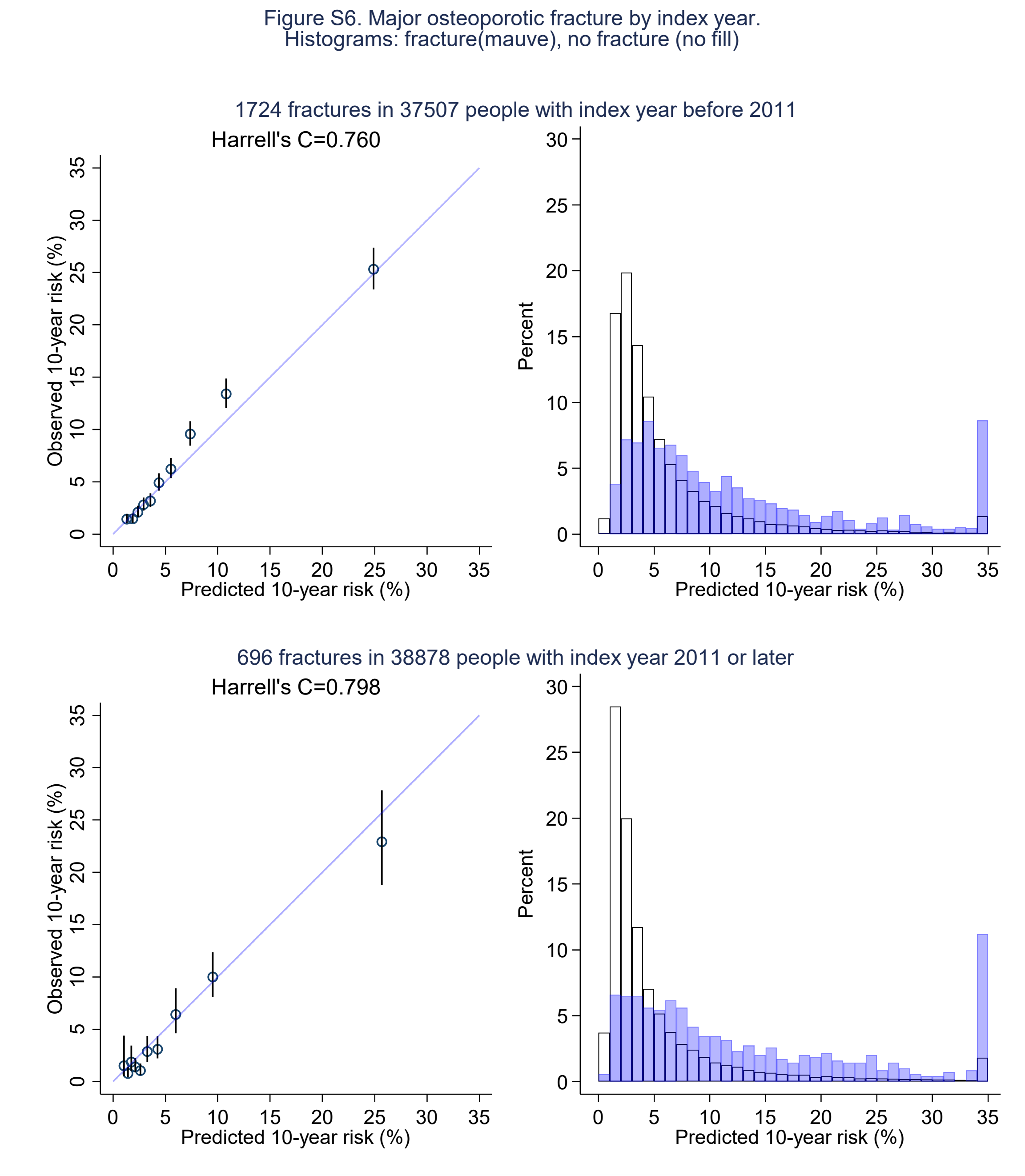


Figure S6 Calibration plots for the performance of the major osteoporotic fracture (MOF) prediction model in the Aurum validation data. Groups are deciles of predicted risk. Validation was done by subgroup of year of index date.

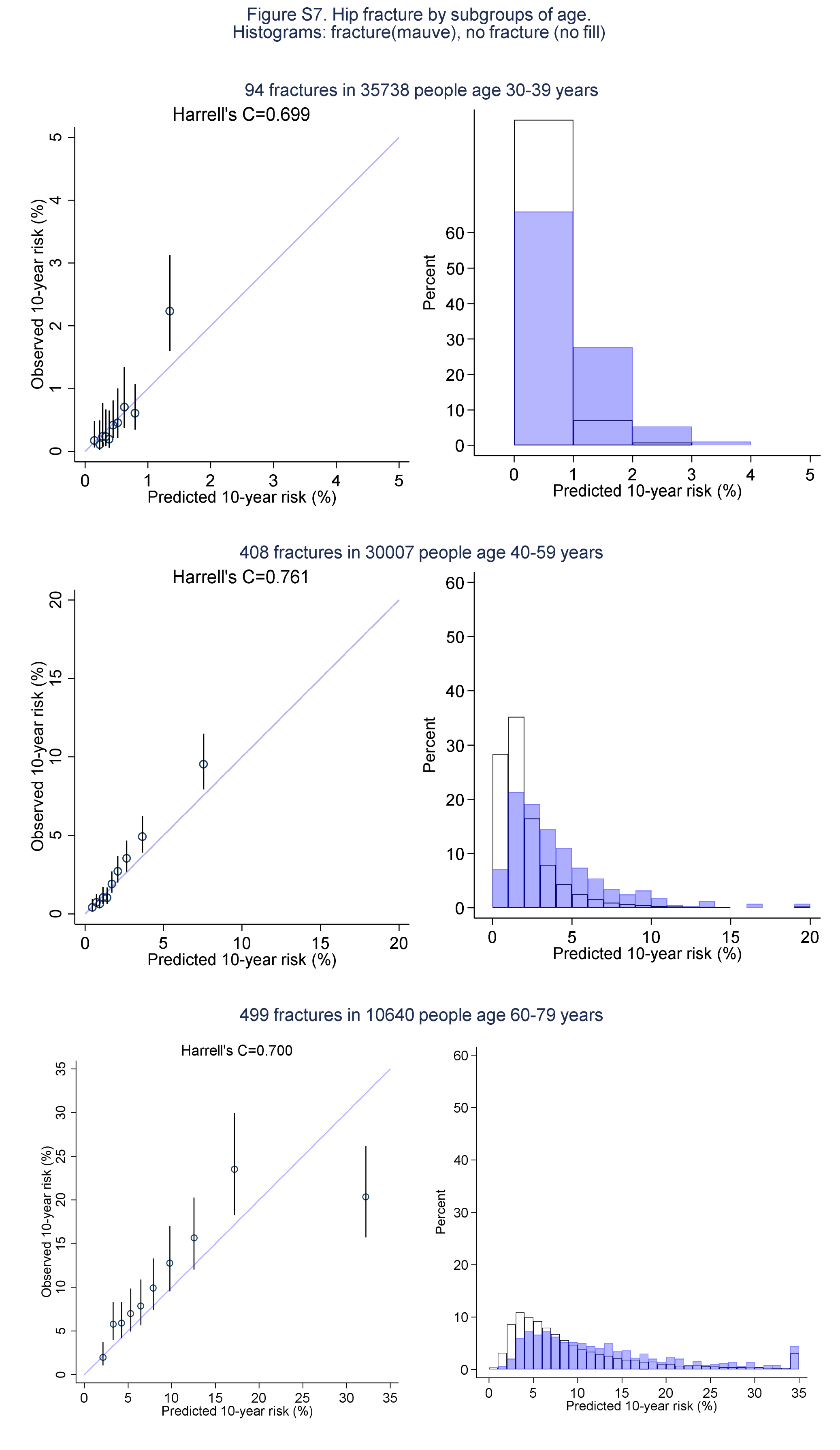


Figure S7 Calibration plots for the performance of the hip fracture prediction model in the Aurum validation data. Groups are deciles of predicted risk. Validation was done by subgroup of age at index.

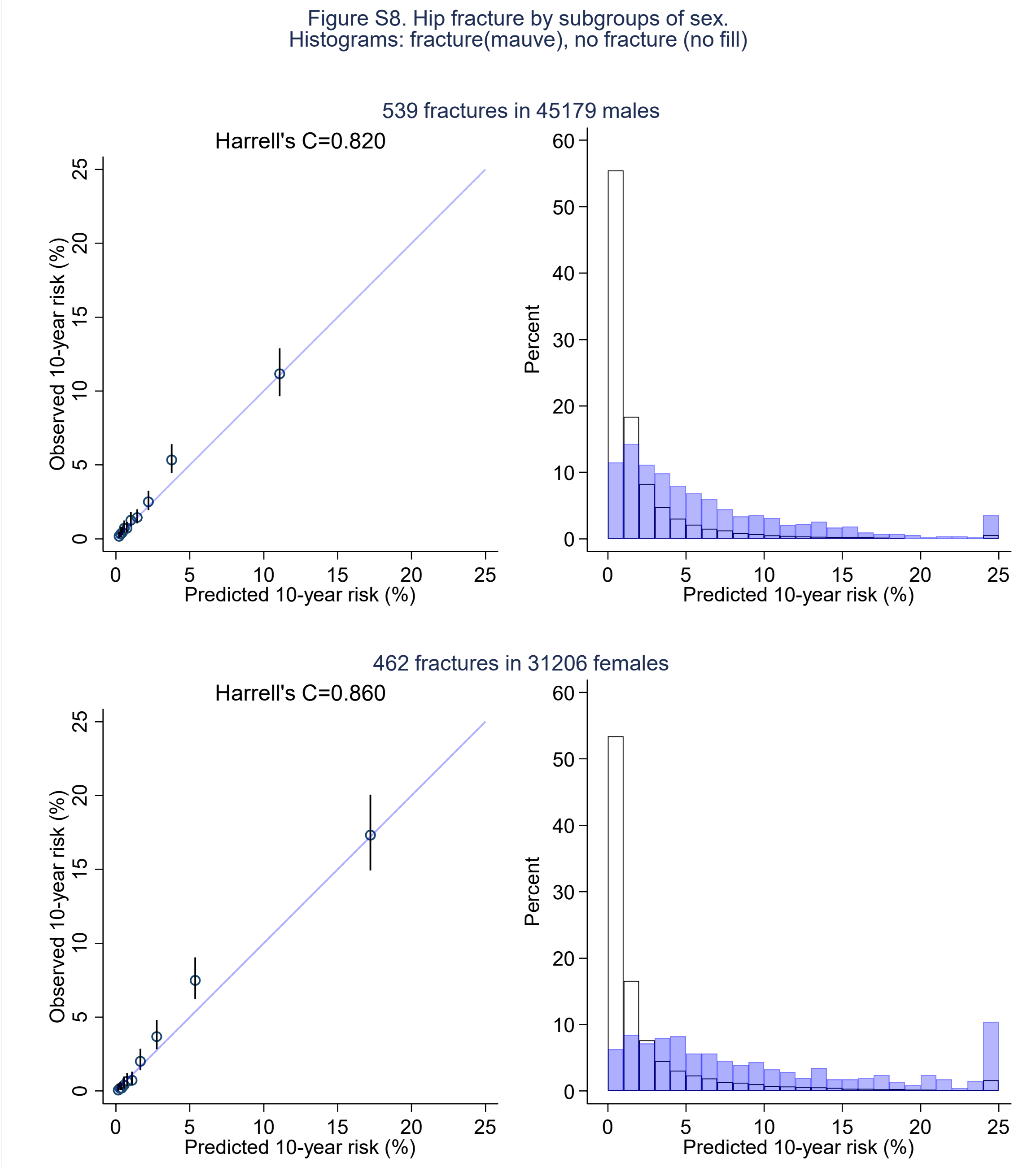


Figure S8 Calibration plots for the performance of the hip fracture prediction model in the Aurum validation data. Groups are deciles of predicted risk. Validation was done by subgroup of sex.

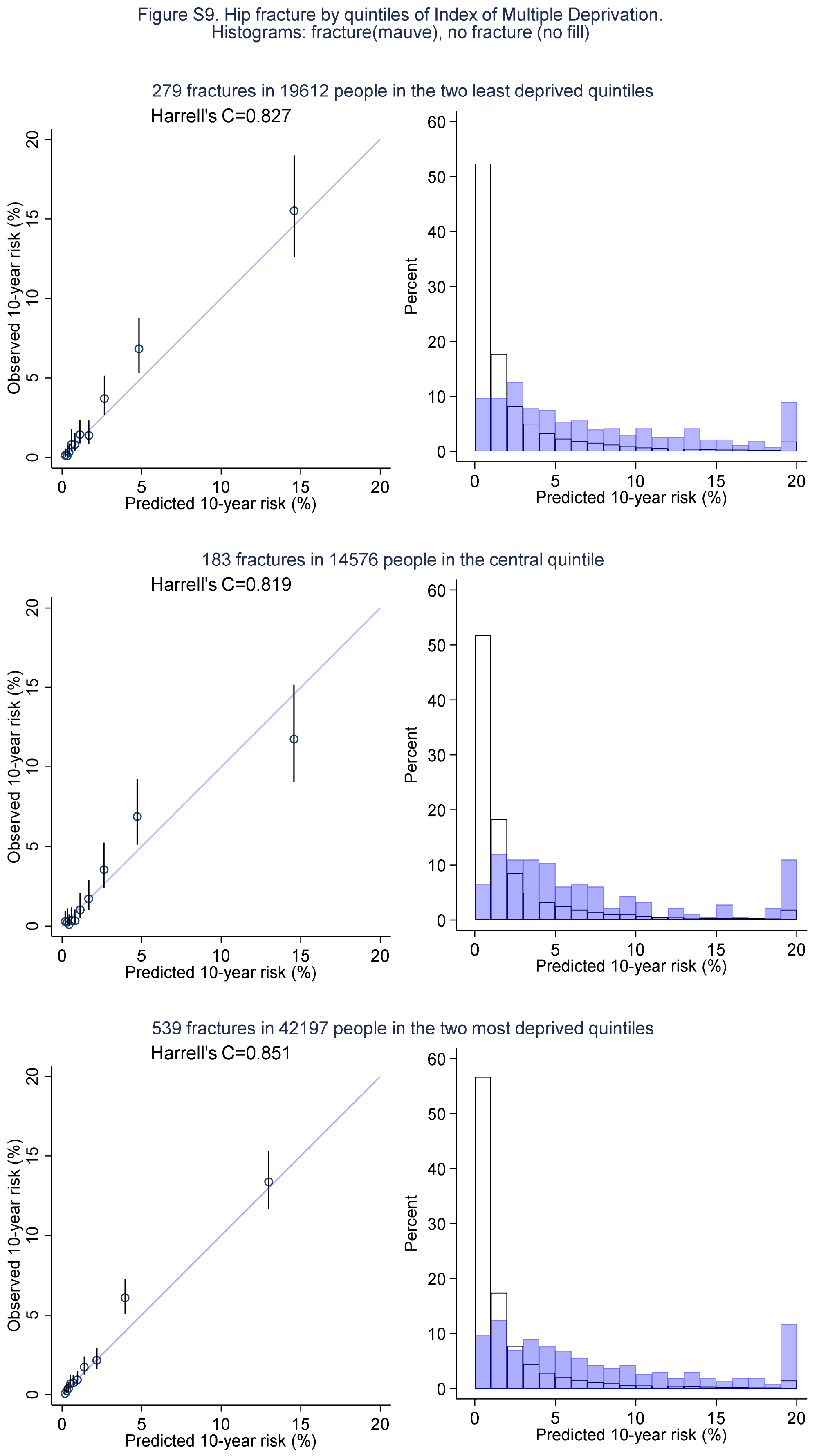


Figure S9 Calibration plots for the performance of the hip fracture prediction model in the Aurum validation data. Groups are deciles of predicted risk. Validation was done by subgroup of Index of Multiple Deprivation.

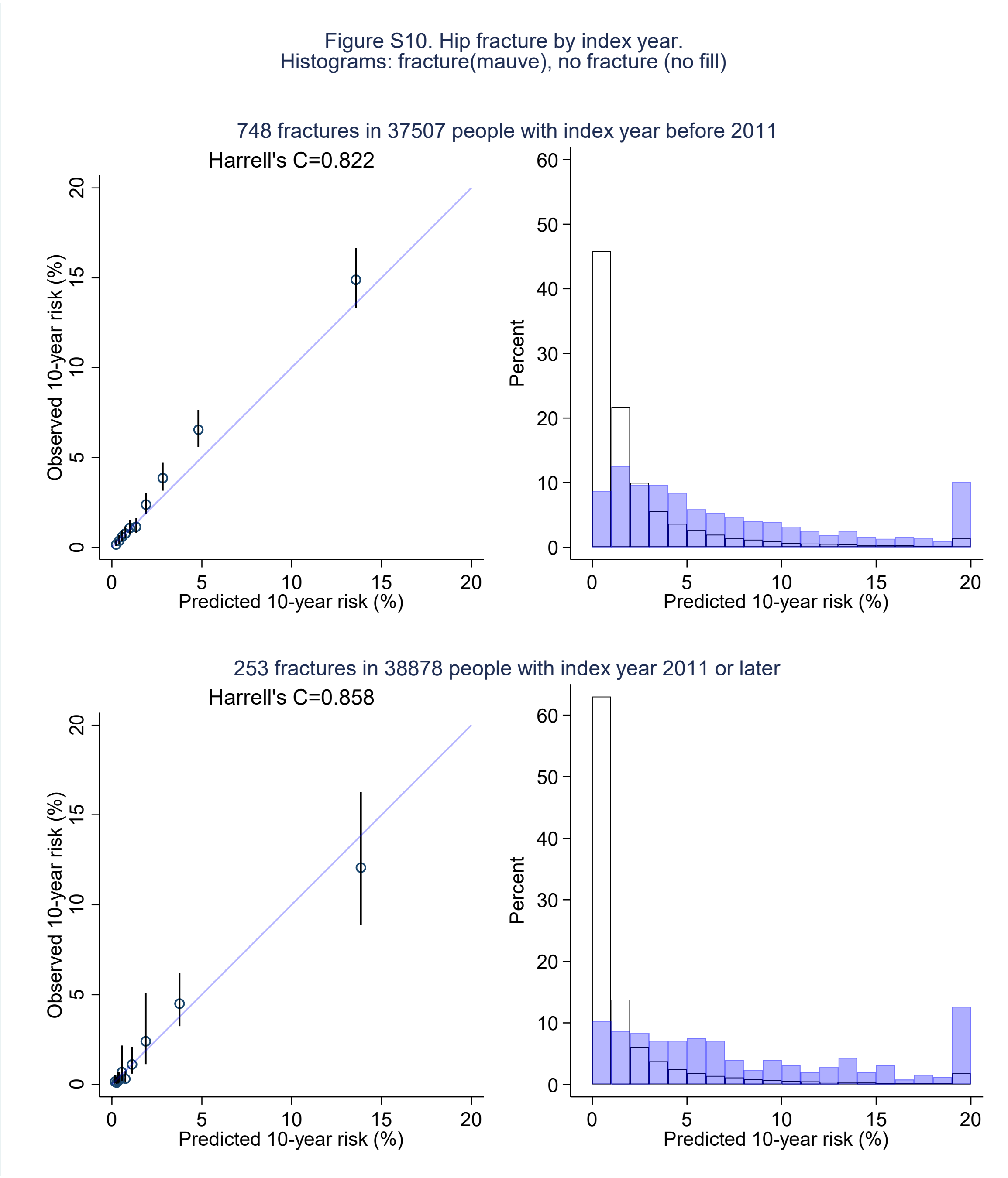


Figure S10 Calibration plots for the performance of the hip fracture prediction model in the Aurum validation data. Groups are deciles of predicted risk. Validation was done by subgroup of year of index date.
